# A novel 5E training model and other methods implemented for rapid readiness at an ad-hoc COVID-19 facility in India

**DOI:** 10.1101/2021.02.23.21251976

**Authors:** Suhail Singh, Anchit Raj Singh, Basant Pathak, TVSVGK Tilak, Aparajita Kumar, IPS Bhatia, Jayraj Hasvi

## Abstract

**Background:** A ad-hoc dedicated COVID-19 hospital was setup in New Delhi, India over a span of 12 days. At this time, new teaching modalities were employed to train the staff. This study aims to identify and quantify the effectiveness of these teaching models in terms of learning which was evaluated using case scenarios before and after the teaching session.

**Objectives:** To assess education methods and models for training of healthcare workers during rapid deployment at an ad-hoc dedicated COVID-19 hospital.

**Methods:** The 5E (Engage, Explore, Explain, Elaborate and Evaluate) teaching modality through peer group teaching methods was utilised in the situation. Statistical analysis was done using Mann Whitney U test.

**Results:** A total of 86 participants (43 doctors and 43 nurses) answered the pre and post test questionnaires. The number of correct responses per question in the pre test (Mean ± SD;49.0± 18.53) vs post test (Mean ± SD;54.40± 15.95) with stratification by the domain of learning was analysed. No significant difference was found in the pre and post test responses.

**Conclusion:** Further studies of this nature will contribute towards assessing efficacy of teaching modalities employed in rapid readiness for future pandemic scenarios.

## Introduction

At the dawn of 21^st^ century, mankind was faced with a daunting challenge, the Coronavirus Disease 2019 (COVID-19) pandemic. The disease was believed to have been originated in the Wuhan province of Chine, in December 2019 (Singhal 2020). The World Health Organisation declared this as a pandemic on 11 March 2020. In the current scenario, with the disease spreading over 220 countries and affecting over 17 million people and a death toll of more than half a million, healthcare facilities have been established throughout the world to tackle the rising numbers (Bedford et al. 2020; Bouadma et al. 2020).

In these desperate times, the healthcare providers have come up with newer means to battle this disease. This includes establishment of ad hoc healthcare facilities built exclusively for COVID-19 positive patients. These include a 1000 bedded temporary COVID-19 hospital that has been setup in the Indian capital, New Delhi over a span of 12 days. The hospital was run by staff flown to Delhi and deployed in this hospital over a span of 2 days. The health care workers (HCWs) of varied experience and specialities, needed innovative teaching modalities to provide them with latest information regarding management of COVID-19 in as little time as possible, while ensuring that adequate working knowledge was communicated. This was attempted using the 5E (Engage, Explore, Explain, Elaborate and Evaluate) teaching modality through peer group teaching methods. Here, we look at the newer teaching methods and models that were used during one such session and their impact on the knowledge and behaviours of the HCWs.

The study aims to identify and quantify the effectiveness of these teaching models in terms of learning which was evaluated using case scenarios before and after the teaching session using web based survey tools.

## Methods

This study reports results of a pre and post teaching cross-sectional online questionnaire conducted in a population of health care workers, which included doctors and nurses deployed in management of COVID-19 infection in a temporary 1000 bedded COVID-19 hospital. Four case scenarios, each with five questions were handed out. The answers were collected on the spot. The questions were divided to assess cognitive and affective domains of learning (eight of the twenty questions were affective and the remaining cognitive). The learning modalities used were 5E and peer group teaching methods.

The study was based in a dedicated 1000 bedded COVID-19 hospital, set up in the national capital of India, Delhi, in view of the surge in number of COVID 19 cases. HCWs involved in direct patient care in this hospital constituted the study population. Our study included a total of 86 participants, including 43 doctors and 43 nurses. Clearance from the Institutional Ethics Committee was taken.

An hour-long session was conducted for both doctors and nurses, before which, an online form tool comprising four case scenarios with five questions each, was attempted the participants. This was used to assess the baseline knowledge of the individuals. The case scenarios and questions were developed by doctors who had prior experience in dealing with COVID-19 cases. This was followed by one-hour teaching session, which was based on the 5E model (Barrow 2006; Açişli et al. 2011). Post the interactive session, the participants were handed out the same questionnaires, to the assess the efficacy of teaching. The clinical scenarios were read out and the nuances of case management were explained by the residents. A flipped classroom session was created, with the participants putting forth their doubts and ideas, which were subsequently dealt with by the residents. Statistical analysis was done using Mann Whitney U test and p values < 0.05 were considered significant.

## Results

A total of 86 participants (43 doctors and 43 nurses) gave consent and answered the pre and post-test questionnaires. The number of participants correctly answering a question increased from a mean of 49 participants in the pre teaching test to a mean of 51 participants in the post teaching module (Table 1). There was increase in the mean number of correct responses per question of both the cognitive and the affective domain (Figures 1-2). However, on statistical analysis no significant difference was found in the means of the pre and post test responses (p value 0.37). On sub group analysis of the responses to the questions of the affective and cognitive domains, there was no statistically significant difference (p value affective domain-0.49, p value cognitive domain-0.27).

**Table 1.** Pre and post test responses stratified by domain of learning

|  | Pre-test number of correct<br>responses per question |  |  | Post-test number of correct<br>responses per question |  |  | P value |
| --- | --- | --- | --- | --- | --- | --- | --- |
|  | Mean | Standard<br>Deviation | Percentage | Mean | Standard<br>Deviation | Percentage |  |
| All | 49.00 | 18.53 | 56.97 | 54.40 | 15.95 | 63.25 | 0.37 |
| Affective<br>domain | 55.75 | 21.06 | 67.73 | 59.75 | 20.24 | 69.48 | 0.49 |
| Cognitive<br>domain | 44.5 | 15.98 | 51.74 | 50.83 | 12.00 | 59.11 | 0.27 |

**Figure 1.**
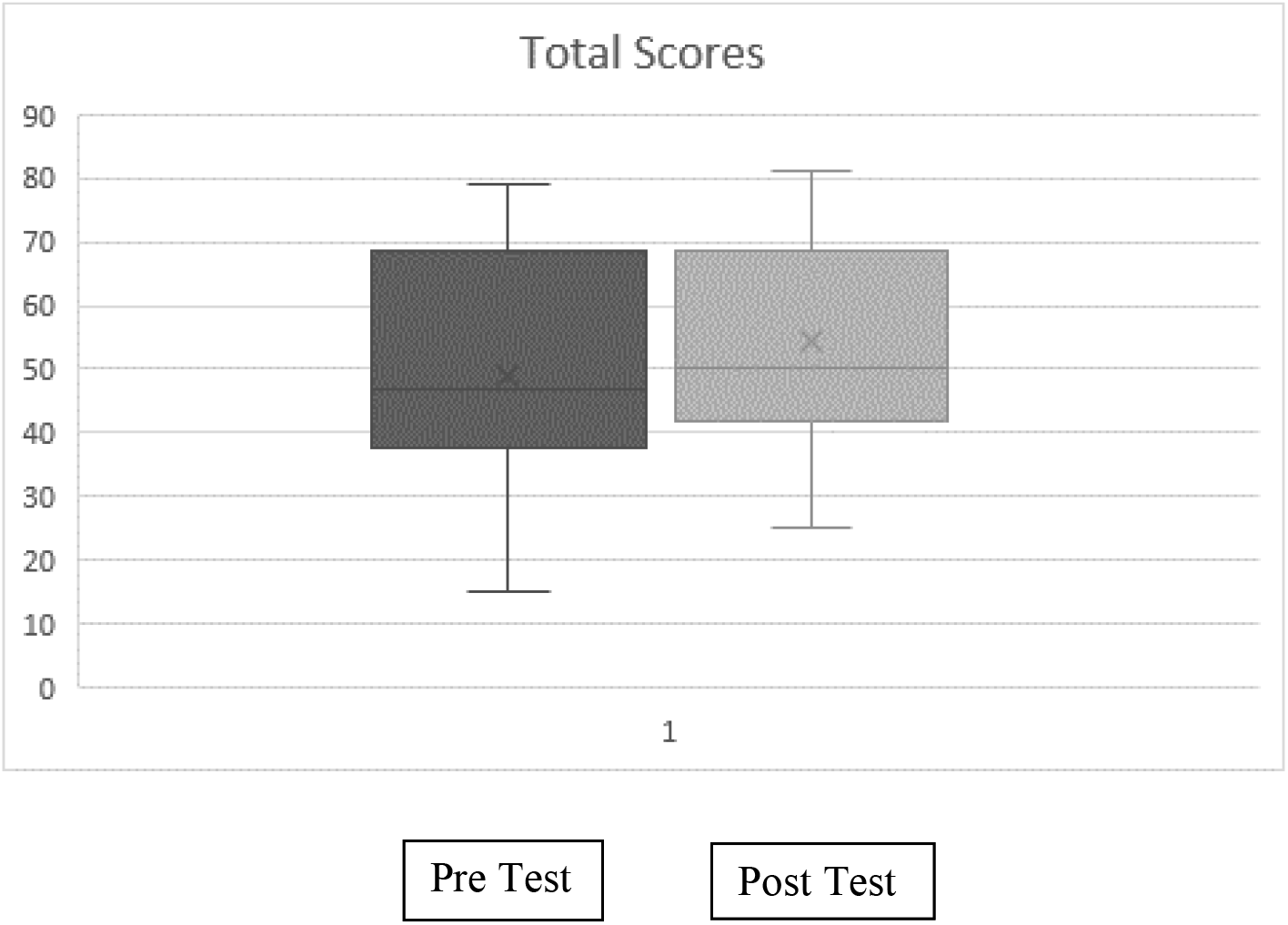
Box and whisker plot of pre and post test number of correct responses for all questions

**Figure 2.**
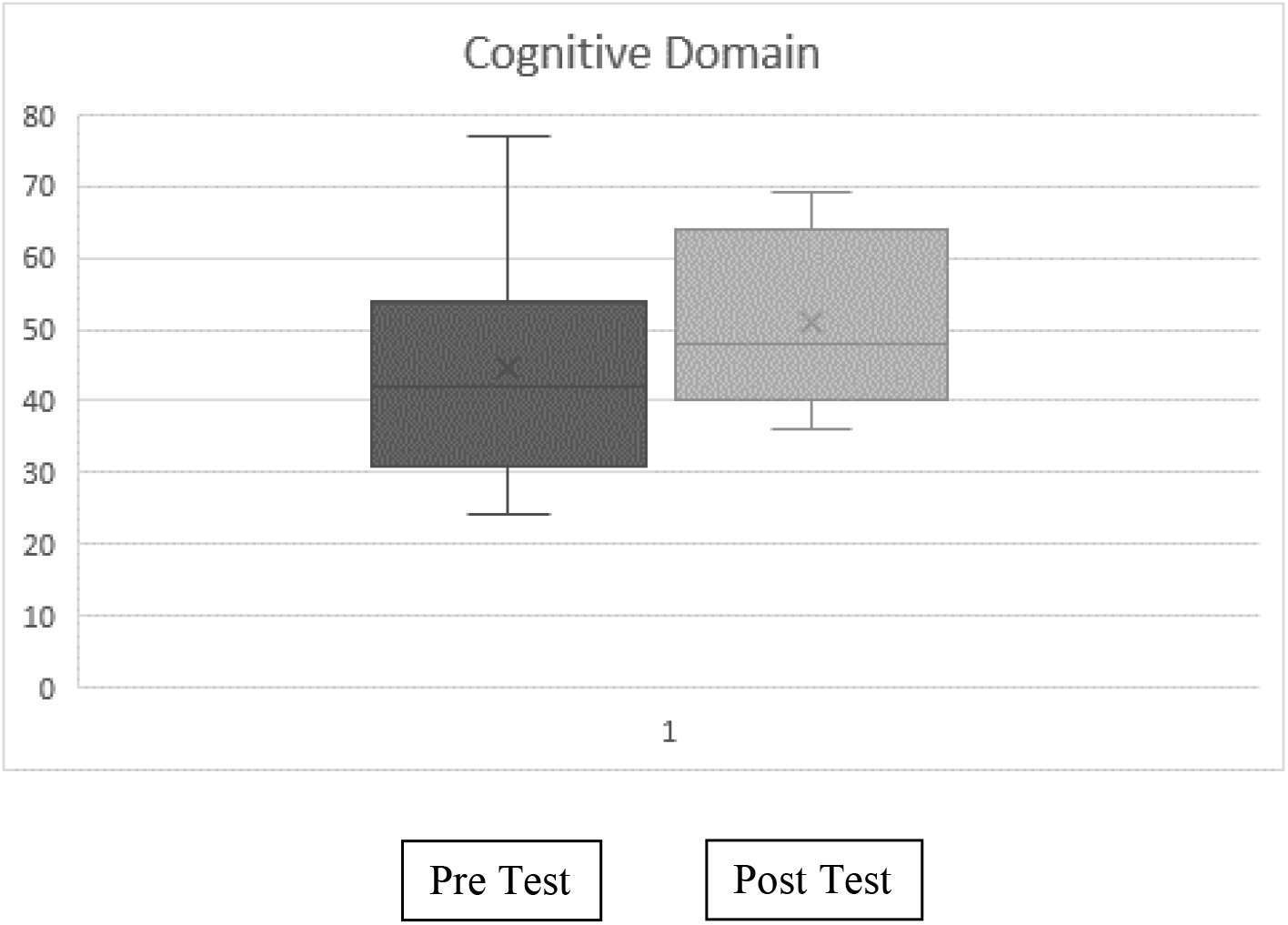
Box and whisker plot of the pre and post test number of correct responses for the questions of the cognitive domain

## Discussion

The COVID-19 pandemic has taken a toll on the healthcare facilities and these testing times have created a strong demand for newer teaching methods for training of healthcare workers, to be ready for deployment in a short span of time. This was a unique setting which offered an opportunity to investigate newer educational practices that could be used for effective manpower training. Our study brings forth the differences in the response of HCWs to newer teaching modalities while being rapidly deployed for the COVID-19 duties.

Our study population included doctors and nurses from across clinical and paraclinical specialities who did not have any significant prior experience in the management of COVID-19 cases. Hence, it was necessary that they be adequately trained before they start manning ICUs to manage active cases. Due to the rapid deployment of workforce, conventional methods of curriculum design and classroom teaching could not be used. In the arena of pedagogy, it has long been recognised that there is a ‘learning cycle’. The students have to go through this cycle in order to inculcate and apply the facts being taught. This idea of the order of events involved learning i.e ‘the learning cycle’ is not new and was first explored by Dewey in the year 1971. Mike Atkin and Robert Karplus in 1962 argued that exploration, term introduction and concept application were the three-key concept of learning (Karplus 2002). The students were made to be interested in the subjects by allowing them to ‘Explore’ the topic, followed by new ‘Term introduction’, which was later built upon by ‘concept application’ in the form of classroom tests and practical exercises. The 5 E model introduced by Bybee and colleagues in the year 2006 is a direct descendent of the Atkin and Karplus method of learning (Bybee 2006). And further classifies the teaching methodology into Engagement, Exploration, Explanation, Elaboration and Evaluation. ‘Engagement’ primarily refers to how a teacher incites inquisitiveness among the participants by assessing their previous knowledge and promoting curiosity through short activities. ‘Exploration’ allows students to ask questions and try and look into the nuances of the topics being taught. The ‘Explain’ phase requires both students and teachers to describe the ideas already learnt during the previous two phases. In ‘Elaboration’ the teachers challenge the limits of the concept understanding and introduce newer ideas related to the subject. Finally, the ‘Evaluate’ phase is when either the students themselves or the instructors assess the knowledge imparted in the previous phases. One important aspect of the 5E learning method is the active student participation. The same was ensured in this scenario. And a structured approach ensured that all the steps were meticulously followed.

Another learning method extensively used in our setting was that of the ‘peer group learning’. Peer group learning basically comprises of students teaching other students about subjects on which they have more knowledge and experience than the others (Dehghani et al. 2014). In this context, the internal medicine residents who are students themselves but had prior experience in dealing with COVID-19 patients were involved both in generating the case scenarios and the questions, and subsequently answering the queries of the participants on this subject. Peer group teaching is known to inculcate more confidence among the participants as it makes the teacher more accessible and inculcates the basic ‘Exploration’ aspect of learning.

The usage of the above-mentioned methods in the setting of rapid deployment of HCWs during the times of COVID-19 is something which has not been explored. Our findings showed an increase in the learning in both the Affective and Cognitive domains. There was no significant difference in the number pre-test and post-test correct responses, even though there was an increase in the absolute number of correct responses, however this could be a result of the small sample size. However more studies on this subject should be undertaken so that an effective training methodology could be developed for HCWs during these trying times.

## Conclusion

The usage of novel methods of teaching for HCWs deployed in COVID-19 hospital shows promise. In this preliminary study, the participants showed positive outcome in terms of the increase in correct responses to an online questionnaire based on case scenarios inculcating concepts and ideas behind management of COVID-19 patients, even though the results are not significant for which we might require better powered studies. The affective and cognitive domains both showed an increase in the absolute number of correct responses. As newer studies and case series are providing us with ever so evolving knowledge about COVID-19, such teaching methods are an important part of the armamentarium of the organisations all over the world to train HCWs even with a time crunch and keep them in the loop with latest findings regarding COVID-19.

## Data Availability

All the Data has been displayed in the original manuscript

## Acknowledgements

Nil

